# Neutralizing Antibody Activity Against SARS-CoV-2 Variants in Gestational Age-Matched Mother-Infant Dyads

**DOI:** 10.1101/2021.12.09.21267557

**Authors:** Yusuke Matsui, Lin Li, Mary Prahl, Arianna G. Cassidy, Nida Ozarslan, Yarden Golan, Veronica J. Gonzalez, Christine Y. Lin, Unurzul Jigmeddagva, Megan A. Chidboy, Mauricio Montano, Taha Y. Taha, Mir M. Khalid, Bharath Sreekumar, Jennifer M. Hayashi, Pei-Yi Chen, G. Renuka Kumar, Lakshmi Warrier, Alan H.B. Wu, Dongli Song, Priya Jegatheesan, Daljeet S. Rai, Balaji Govindaswami, Jordan Needens, Monica Rincon, Leslie Myatt, Ifeyinwa V. Asiodu, Valerie J. Flaherman, Yalda Afshar, Vanessa L. Jacoby, Amy P. Murtha, Joshua F. Robinson, Melanie Ott, Warner C. Greene, Stephanie L. Gaw

## Abstract

Pregnancy confers unique immune responses to infection and vaccination across gestation. To date, there is limited data comparing vaccine versus infection-induced nAb to COVID-19 variants in mothers during pregnancy. We analyzed paired maternal and cord plasma samples from 60 pregnant individuals. Thirty women vaccinated with mRNA vaccines were matched with 30 naturally infected women by gestational age of exposure. Neutralization activity against the five SARS-CoV-2 Spike sequences was measured by a SARS-CoV-2 pseudotyped Spike virion assay. Effective nAbs against SARS-CoV-2 were present in maternal and cord plasma after both infection and vaccination. Compared to wild type or Alpha variant Spike, these nAbs were less effective against the Kappa, Delta, and Mu Spike variants. Vaccination during the third trimester induced higher nAb levels at delivery than infection during the third trimester. In contrast, vaccine-induced nAb levels were lower at the time of delivery compared to infection during the first trimester. The transfer ratio (cord nAb level/maternal nAb level) was greatest in mothers vaccinated in the second trimester. SARS-CoV-2 vaccination or infection in pregnancy elicit effective nAbs with differing neutralization kinetics that is impacted by gestational time of exposure. Vaccine induced neutralizing activity was reduced against the Delta, Mu, and Kappa variants.

**Graphic abstract:** 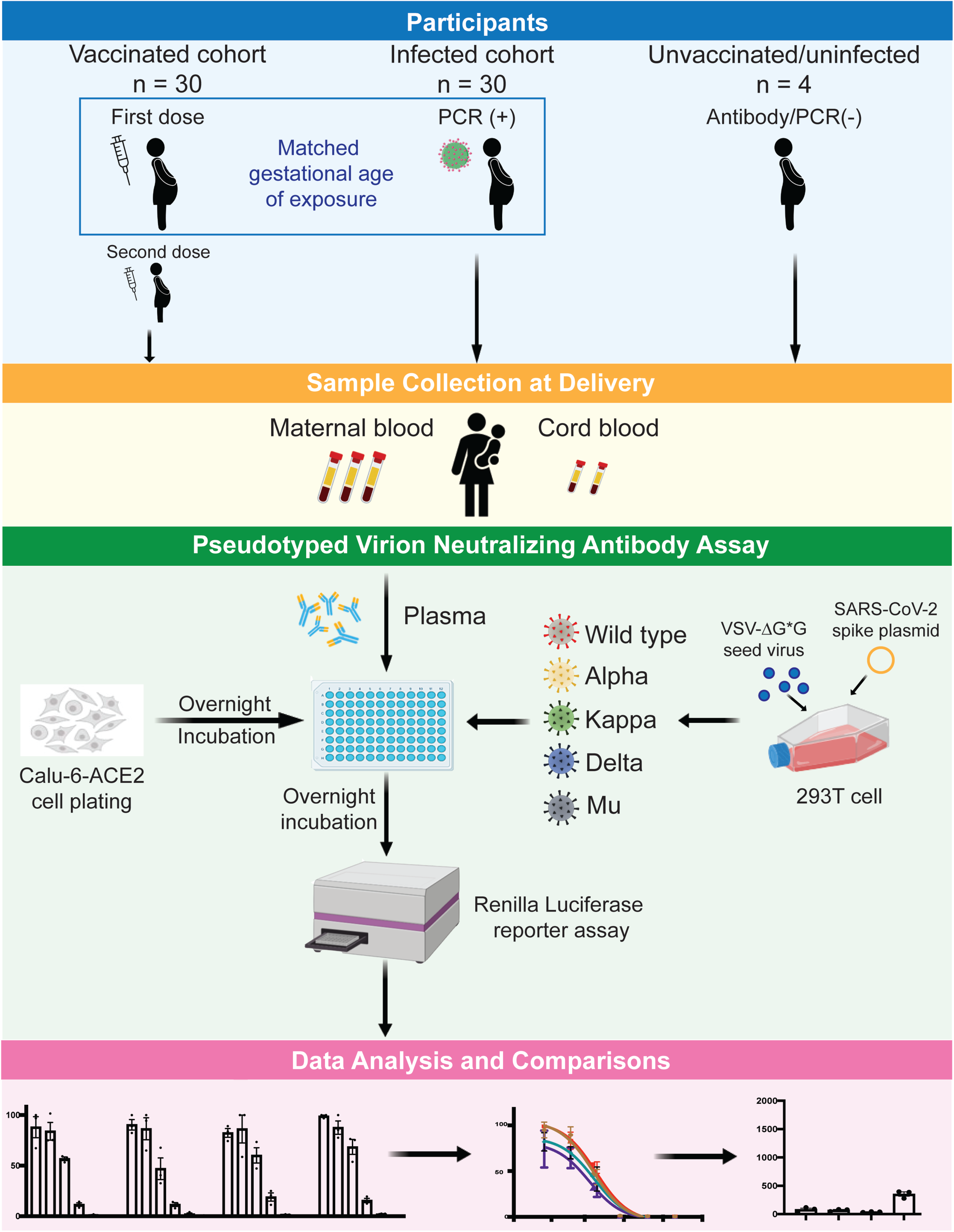

## Introduction

Pregnant women are identified as an at-risk population for severe coronavirus disease 2019 (COVID-19) with increased rate of ICU admission, invasive mechanical ventilation, and death (1–4). COVID-19 infection has exacerbated long standing perinatal inequities among communities that already experienced higher rates of maternal mortality and morbidity and poor infant outcomes (5, 6). All leading professional and public health organizations have strongly recommended that pregnant individuals be vaccinated against severe acute respiratory syndrome coronavirus 2 (SARS-CoV-2) (7). Although pregnant individuals were excluded from the first clinical trials of COVID-19 vaccines, a growing body of evidence indicates that fully vaccinated pregnant women have a significantly lower risk of SARS-CoV-2 infection due to the generation of robust humoral and cellular immunity (8–11). Furthermore, maternal IgG antibodies induced after vaccination and natural infection can be detected in umbilical cord blood of newborns at birth (10, 12), these antibodies may protect newborns from SARS-CoV-2 infection in early life.

Two FDA-authorized mRNA vaccines that encode the SARS-CoV-2 spike protein, BNT162b2 (Pfizer-BioNTech) and mRNA-1273 (Moderna), exhibit greater than 90% efficacy against cases of COVID-19 up to at least 4 months (13). However, genetic mutations in the Spike protein change the transmissibility, and sensitivity to neutralizing antibodies (nAbs). The WHO designates the Alpha (B.1.1.7), Beta (B.1.351, Gamma (P.1), Delta (B.1.617.2) and Omicron (B.1.1.529) variants as variants of concern (VOC) to help track SARS-CoV-2 genetic lineages. The Delta variant is highly contagious and led to the 2021 resurgence of COVID-19 worldwide. Early data suggests sera from fully vaccinated or convalescent non-pregnant individuals display reduced neutralizing activity against the Delta variant compared to the Alpha variant (14). The Kappa variant (B.1.617.1) originated from the same lineage as the Delta and is categorized as variants under monitoring (VUM) based on the epidemiological significance (15). The newly recognized Mu variant of interest (B.1.621) is described as 10.6-fold and 9.1-fold more resistant to sera from COVID convalescents or BNT162b2-vaccinated individuals respectively compared to the ancestral wild type virus (16).

Robust humoral response elicited by SARS-CoV-2 infection and vaccination, and efficient IgG transfer in pregnancy have been widely reported (10, 17–21). Previous studies indicate that the receptor-binding domain (RBD) of the SARS-CoV-2 Spike protein contains multiple conformational neutralizing epitopes (22). These include sites where single amino acid mutations including N501 (Alpha and Delta), L452 (Kappa and Delta), and E484 (Kappa and Mu) compromise antibody-mediated neutralization and/or transmission (**Figure 1**). The N501Y mutation is the only mutation in the interface between the RBD of the Alpha variant and does not have strong impact on the activity of most nAbs induced by natural infection and vaccination but improves viral transmission (23, 24). The Kappa and Delta variants possess the L452R mutation, which modestly affects nAb binding affinity (25). The Kappa variant possesses the E484Q mutation, which reduces sensitivity to vaccine-elicited nAb, but the L452R and E484Q mutation are not synergistic for loss of sensitivity to neutralizing antibodies (26). The Mu variant harbors the E484K which has been shown to significantly reduce sensitivity to the nAbs (27, 28). Furthermore, the Delta variant has the specific T478K mutation, which has been shown to improve viral interaction with ACE2, but there is little knowledge about its potential role in resistance to neutralization by antibodies (29). The Mu variant also has the specific R346K mutation, which may affect the sensitivity of nAb (30).

**Figure 1.**
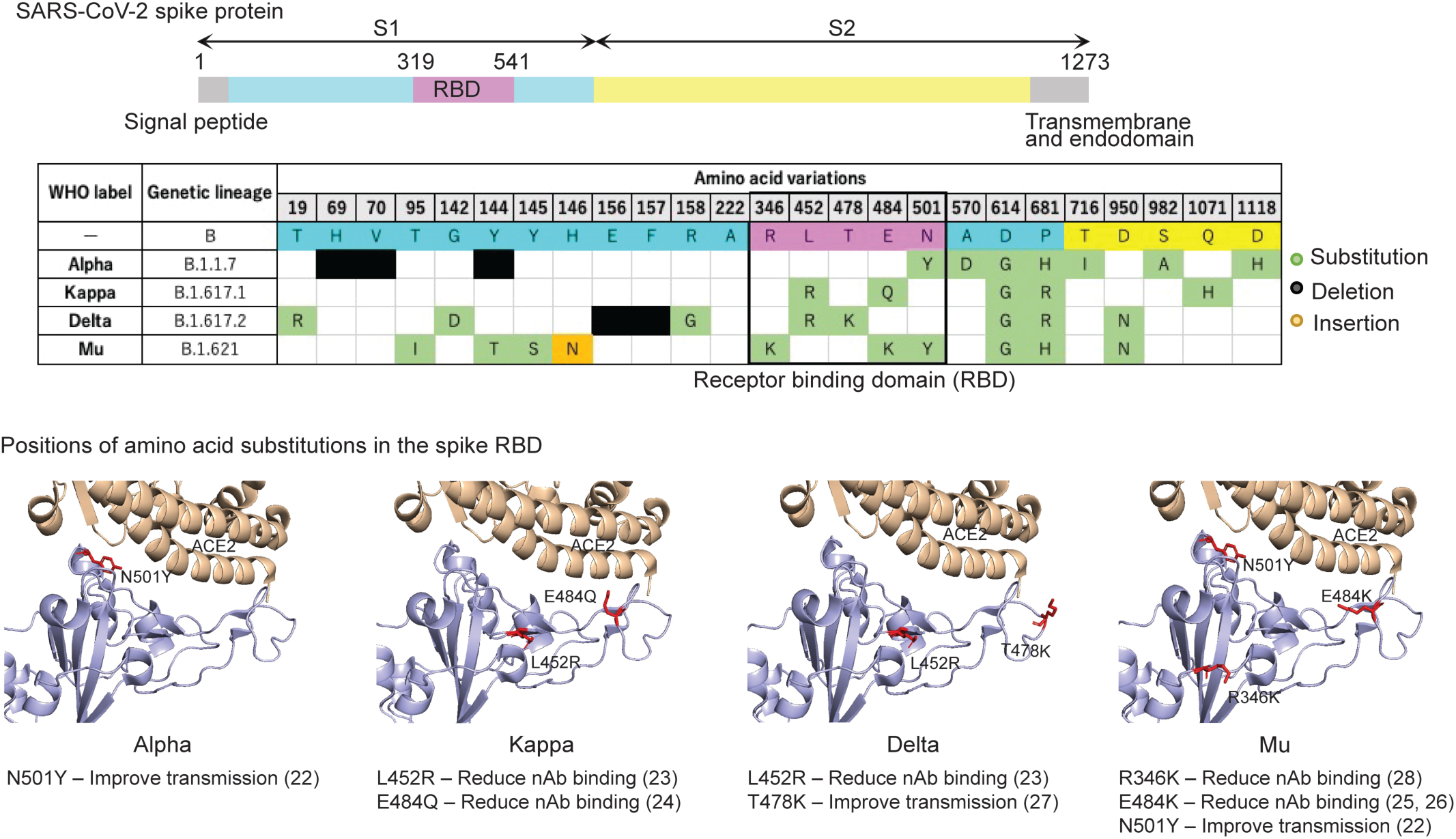
Positions of mutated amino acid residues in the Spike proteins from four variants. The residues L452R, T478K, R346K, E484K, and N501Y are labeled in red. The crystal structure of SARS-CoV-2 spike RBD and ACE2 (PDB: 6M0J) (64) was used as the template. All structure figures were generated with PyMol (65).

Existing studies indicated that maternal IgG production and maternal-fetal transfer might be influenced by timing of exposure, fetal sex or antibody glycosylation profiles (11, 31, 32). However, the neutralizing activity against different SARS-CoV-2 variants during pregnancy and the transplacental transfer efficacy over gestation remains understudied. In this study, we assessed nAb activities against five strains of SARS-CoV-2, including the wild-type (WT) (Wuhan-Hu-1), Alpha, Kappa, Delta, and Mu variants in paired maternal and infant cord blood plasma samples collected at the time of delivery. To understand the impact of timing during pregnancy on the development of maternal-fetal humoral immunity, we compared two cohorts that were matched by gestational age of exposure. Using samples from vaccinated versus infected pregnant individuals, we compared and contrasted the nAbs elicited by mRNA vaccine compared with natural infection, their efficacy against newly emerging variants, impact of the timing of vaccination/infection on maternal and neonatal protection, and clinical correlates of nAb production.

## Results

### Participant characteristics

Sixty-four maternal-fetal dyads were included in this study (**Table 1**). Among the 30 vaccinated pregnant women, 5 (16.7%) received the first vaccine dose in the first trimester (0 to 13 weeks and 6 days), 12 (40%) in the second (14 weeks to 27 weeks and 6 days), and 13 (43.3%) in the third (28 weeks to 40 weeks and 6 days). In the matched 30 naturally infected pregnant women, 5 (16.7%) had the first positive PCR result in the first trimester, 13 (43.3%) in the second, and 12 (40%) in the third. Maternal age was significantly higher in the vaccinated group as compared with the infected group (36 vs. 28, *P* < 0.0001). The participants were from all racial or ethnic groups; however, 70% (21/30) vaccinated participants were white and 73.3% (22/30) naturally infected participants were Hispanic/Latina (*P* < 0.0001). Participants delivered at a median of 91 days (IQR, 60-140 days) after the first dose of vaccine from vaccinated mothers (30 dyads), and 92 days (IQR, 53-139 days) after the earliest positive test from infected mothers (30 dyads) (*P* = 0.56). Individuals with infection during pregnancy were more likely to be obese pre-delivery compared to vaccinated group (median BMI, 32.2 vs. 23.5, *P* < 0.0001). There were no differences between groups with respect to gestational age at delivery or fetal sex. Among participants infected with SARS-CoV-2, 7 (23.3%) were asymptomatic, 20 (67.7%) experienced mild disease and 3 (10%) experienced severe disease (defined as having evidence of COVID-19 pneumonia, hypoxia (O_2_ saturation <94%), and/or need for intensive care). Four dyads who were unvaccinated with no evidence of prior infection by serology recruited early in the pandemic served as negative controls.

**Table 1.** Characteristics of pregnant women receiving mRNA COVID-19 vaccine and testing positive for SARS-CoV-2

|  | Vaccinated<sup>a</sup><br>(n=30) | Naturally infected<sup>a</sup><br>(n=30) | Unvaccinated/uninfected<br>(n=4) | *P* value |
| --- | --- | --- | --- | --- |
| Age (years) | 36 (33-38) | 28 (24-36) | 32 (28-35) | < 0.0001 |
| < 35 | 12 (40.0) | 23 (76.7) | 3 (75.0) |  |
| ≥ 35 | 18 (60.0) | 7 (23.3) | 1 (25.0) |  |
| Race/Ethnicity |  |  |  | < 0.0001 |
| White | 21 (70.0) | 2 (6.7) | 3 (75.0) |  |
| Black | 0 | 2 (6.7) | 0 |  |
| Asian | 3 (10.0) | 3 (10.0) | 0 |  |
| Hispanic/Latina | 2 (6.7) | 22 (73.3) | 1 (25.0) |  |
| Other<sup>b</sup> | 4 (13.3) | 1 (3.3) | 0 |  |
| GA at exposure<sup>c</sup> (weeks) |  |  | N/A | 0.96 |
| First trimester<sup>d</sup> | 5 (16.7) | 5 (16.7) | N/A |  |
| Second trimester<sup>d</sup> | 12 (40.0) | 13 (43.3) | N/A |  |
| Third trimester<sup>d</sup> | 13 (43.3) | 12 (40.0) | N/A |  |
| GA at delivery (weeks) | 39.1 (37.7-39.6) | 39.1 (38.7-39.3) | 38.9 (37.5-40.0) | 0.64 |
| Preterm | 3 (10.0) | 2 (6.7) | 0 |  |
| Term | 27 (90.0) | 28 (93.3) | 4 (100.0) |  |
| Fetal sex |  |  |  | 0.80 |
| Male | 14 (46.7) | 13 (43.3) | 4 (100.0) |  |
| Female | 16 (53.3) | 17 (56.6) | 0 |  |
| Maternal BMI<sup>e</sup> | 23.5 (22.0-27.3) | 32.2 (28.8 – 35.6) |  | < 0.0001 |
| 18.5 - 24.9 | 18 | 2 | N/A |  |
| 25 - 29.9 | 7 | 8 | N/A |  |
| ≥ 30 | 5 | 19 | N/A |  |
| Days from exposure<sup>c</sup> to delivery | 91 (60-140) | 92 (53-139) | N/A | 0.56 |
| Disease Severity |  |  |  | N/A |
| Asymptomatic | N/A | 7 (23.3) | N/A |  |
| Mild | N/A | 20 (67.7) | N/A |  |
| Severe | N/A | 3 (10.0) | N/A |  |
| Vaccine |  |  |  | N/A |
| mRNA-1273 | 13 (43.3) | N/A | N/A |  |
| BNT162b2 | 17 (56.7) | N/A | N/A |  |
| Days after second dose |  |  |  | N/A |
| ≥14 and < 28 | 3 (10.0) | N/A | N/A |  |
| ≥ 28 | 27 (90.0) | N/A | N/A |  |

Data are presented as n (%) or median (interquartile range) unless otherwise indicated.
GA, gestational age; BMI, body mass index.
<sup>a</sup>Participants were selected to match based on the approximated gestational age of first-dose vaccine and positive test
<sup>b</sup>Other includes multiracial and unknown
<sup>c</sup>Exposure is defined as date of first dose vaccine or SARS-CoV-2 PCR positive.
<sup>d</sup>The trimesters are defined according to the American College of Obstetricians and Gynecologists.
<sup>e</sup>Maternal BMI was measured at delivery.

### Maternal and cord neutralizing antibodies elicited by vaccination and infection

We assessed the ability of paired maternal and infant cord plasma to neutralize entry of SARS-CoV-2 pseudotyped virions into Calu-6-ACE2 target cells and compare the neutralization potency of plasma from the vaccinated and infected participants. No neutralizing activity was detected in the four unvaccinated, uninfected maternal and infant dyad samples (**Supplemental Figure 1A).** The composite median NT50 titers against all examined strains was higher in cord blood from vaccinated mothers than those from infected mothers (202 vs. 104, *P* < 0.0001), but not in maternal blood (128 vs. 120, *P* = 0.12) (**Figure 2A**).

**Figure 2.**
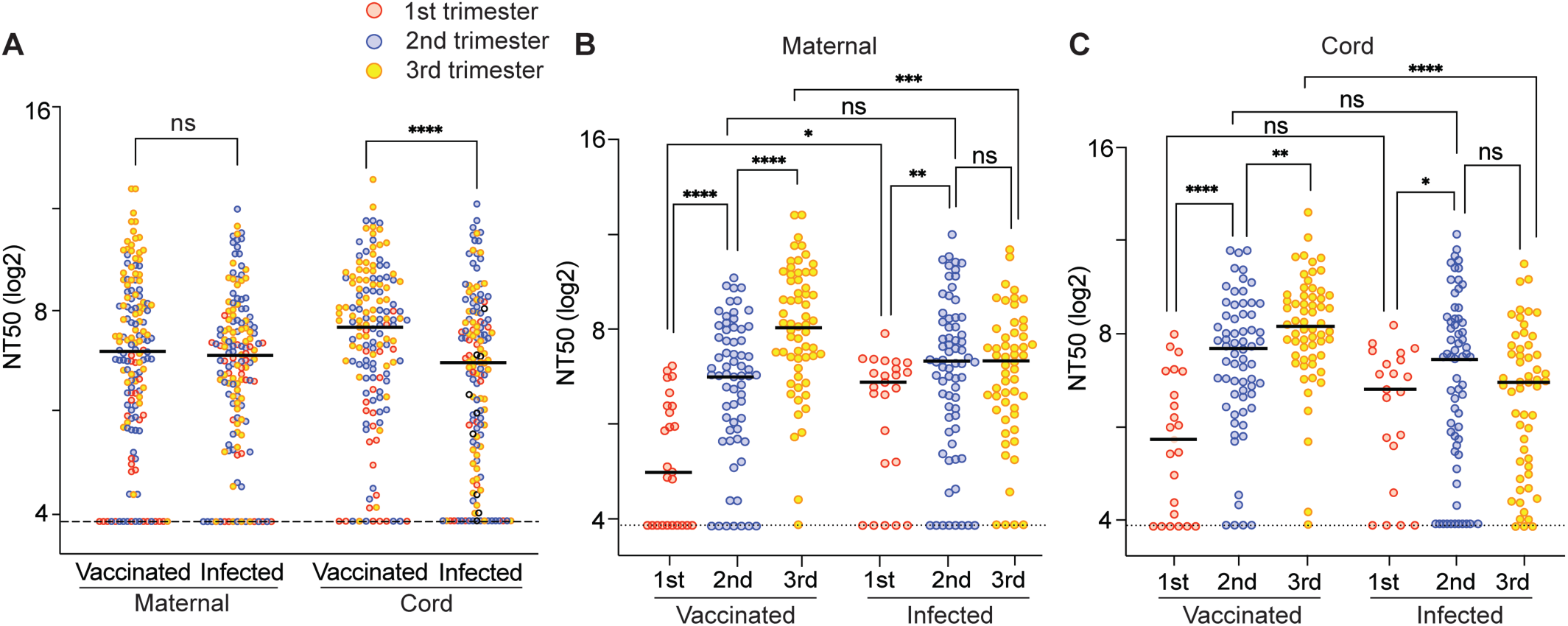
Neutralizing activity of maternal and cord plasma samples from vaccination cohort and infection cohort. (**A**) Comparison of the composite median NT50 titers against all examined strains between vaccinated and infected group. The dot plots show NT50 values in maternal plasma and cord plasma. The dotted line indicates the cut-off threshold of this assay. (**B, D**) NT50 values of the maternal and cord blood were compared separately. Black bars represent the median of NT50 values. \**P* < 0.05; \*\**P* < 0.01; \*\*\**P* < 0.001; \*\*\*\**P* < 0.0001; ns, not significant (Mann-Whitney test). Trimester of exposure is indicated by color (red, first trimester; blue, second trimester; yellow, third trimester).

The total amount of neutralizing activity detected in maternal (**Figure 2B**) and cord (**Figure 2C**) plasma collected at the time of delivery increased with vaccinations later in pregnancy, with the highest maternal and cord NT50 values with maternal vaccinations in the third trimester. In the infected cohort, maternal (**Figure 2B**) and cord (**Figure 2C**) NT50s were lowest in the first trimester compared to second and third trimester infections. Maternal and cord nAb activities at delivery were higher after third trimester vaccination compared to third trimester infection. In first trimester exposures, maternal nAbs were lower at the time of delivery after vaccination than after infection, while there was no difference in cord nAbs after vaccination versus infection. The trend of higher maternal and cord nAb activities after vaccination compared to infection was seen in all five strains in the third trimester. (**Supplemental Figure 1B and 1C**). These results suggest vaccination elicit higher titers of nAb in pregnant individuals within the first few weeks than infection but waned gradually throughout pregnancy. In contrast, infection-elicited antibodies remained relatively stable level until delivery in maternal and cord blood.

### Neutralizing activities against four variants in pregnancy

Emerging variants with mutations in the SARS-CoV-2 RBD raise concern for the efficacy of vaccine-induced and natural immunity, but the nAb activity against these variants in pregnancy is unknown. Here, we analyzed the nAb activities against the Spike proteins from the Alpha, Kappa and Delta and Mu variants in pregnant individuals. In the vaccinated cohort, maternal and cord nAb against the Kappa Spike variant was reduced by 1.7-fold (*P* < 0.0001) and 1.7-fold (*P* < 0.0001), respectively, compared to the WT. Anti-Delta variant was reduced by 2.9-fold (*P* < 0.0001) and 1.8-fold (*P* = 0.0032), in maternal and cord blood, respectively. The Mu variant showed the greatest resistance to nAb inhibition, with maternal and cord plasma showing 4.9-fold (*P* < 0.0001) and 3.0-fold (*P* < 0.0001) reductions in NT50 values, respectively (**Figure 3A**). Neutralizing antibody activity against the Alpha variant was not significantly different from that of the WT. Five of seventeen (29%) women who received BNT162b2 did not demonstrate nAb activity for at least one strain, and three women of thirteen (23%) who received mRNA-1273 did not have detectable nAb activity for at least one strain in maternal blood. There was no difference for the median NT50 values between the two vaccines in maternal or infant cord blood.

**Figure 3.**
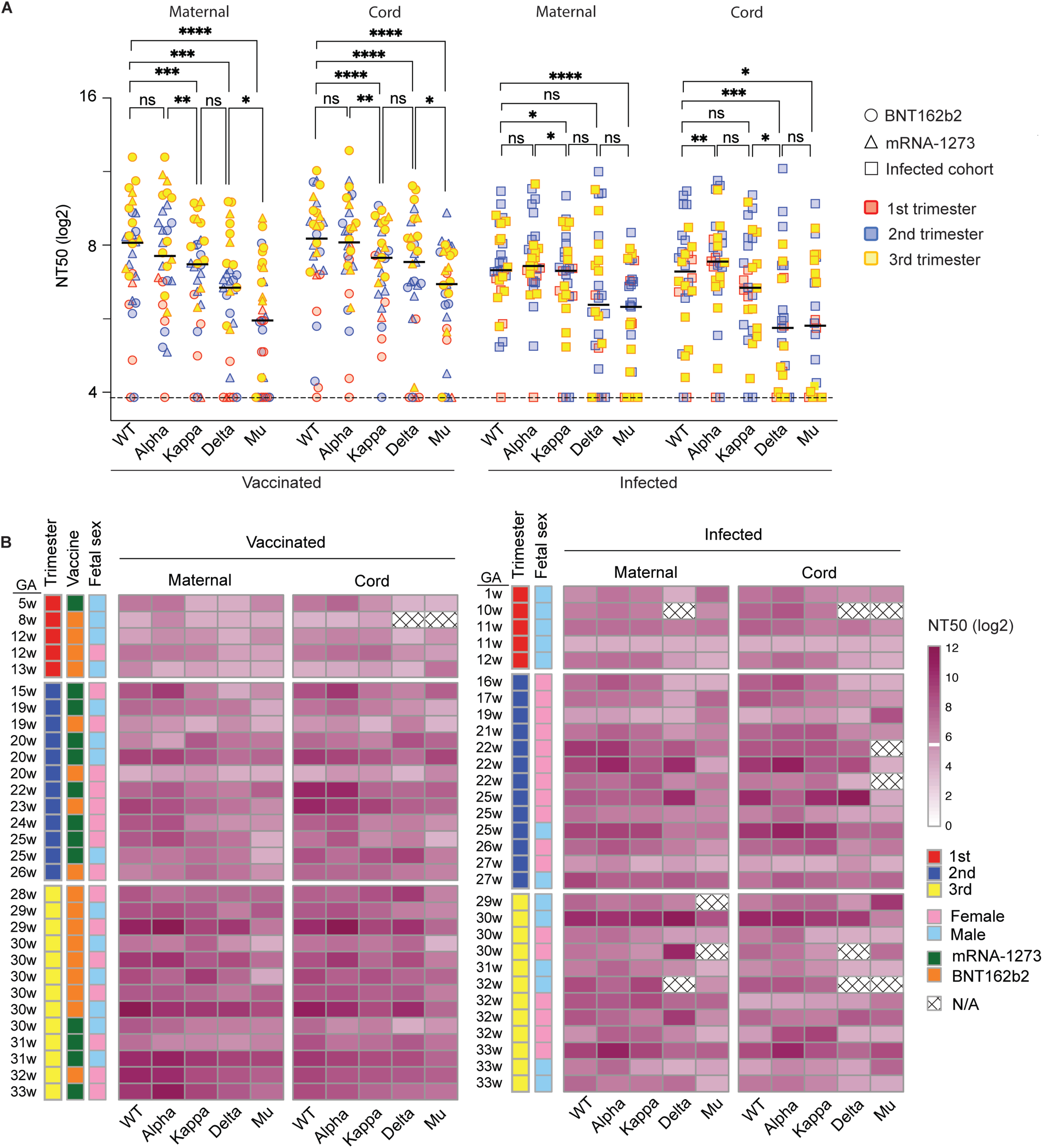
SARS-CoV-2 Neutralizing Antibody Activities in Plasma from Vaccinated and Infected Pregnant Women and Infant Cord Blood Dyads. (**A**) Among a vaccinated cohort and an infected cohort, we matched the timing of the first vaccination to the timing of the infection to be the same. Neutralization assays were performed by exposing Calu-6-ACE2 cells with pseudotyped virions displaying the WT SARS-CoV-2 spike, Alpha, Kappa, Delta, or Mu) to serial dilutions of plasma. 50% neutralization (NT50) values of the plasma samples were defined as the sample dilution at which a 50% reduction in relative light units was observed relative to the average of the virus control wells. Triplicates were performed for each tested serum dilution. Black bars represent the median NT50 values. (**B**) Paired maternal and cord NT50 titers against SARS-CoV-2 strains of each vaccinated and infected patient, ordered by increasing gestational age, grouped by trimester. Patient characteristics are indicated by color (Trimester of exposure: red, first trimester; blue, second trimester; yellow, third trimester. Vaccine type: green, mRNA-1273; orange, BNT162b2. Fetal sex: pink, female; blue, male). N/A, samples are not enough. \**P* < 0.05; \*\**P* < 0.01; \*\*\**P* < 0.001; \*\*\*\**P* < 0.0001; ns, not significant (Wilcoxon signed rank test).

Among the infected cohort, maternal and cord nAb against the Kappa variant was reduced by 1.1-fold (*P* = 0.032) and 1.5-fold (*P* = 0.170), respectively, compared to the WT. Activity against the Delta variant was reduced by 2.1-fold (*P* = 0.120) and 3.1-fold (*P* < 0.0001), and that against the Mu variant was reduced by 2.5-fold (*P* < 0.0001) and 3.0-fold (*P* = 0.036), in maternal and cord blood, respectively (**Figure 3A**). NAb activity against the Alpha variant was not significantly different from that against the WT in maternal plasma but was 1.1-fold higher (*P* = 0.007) than that for the WT in cord plasma. Nine of thirty infected women (30%) did not exhibit nAb activities against at least one of the five strains tested.

Next, we examined individual variability in nAb response across the five strains in maternal and cord blood in the context of exposure type (vaccination versus infection) and trimesters of exposure (**Figure 3B**). We did not observe a significant difference in the variance in nAb response on an individual level between vaccinated and infected cohorts in maternal (*P* = 0.8123) or cord (*P* = 0.5257) blood (**Supplemental Figure 2A and 2B**), or across trimesters of both vaccination and infection cohorts (maternal, *P* = 0.1328; cord, *P* = 0.3694) (**Supplemental Figure 2C and 2D**). In addition, we did not observe a significant interaction between vaccinated/infected cohort and trimester of exposure (maternal, *P* = 0.1722; cord, *P* = 0.3608). Although each participant had variable nAb response to different SARS-CoV-2 strains, neither vaccination/infection cohort nor trimester of exposure significantly contributed to this variance.

### Transplacental transfer of maternal neutralizing antibodies

Reports have shown that maternal to fetal transfer of IgG antibodies was effective and was impacted by the timing of SARS-CoV-2 infection or vaccination (31–33), but the transfer efficiency of nAb induced from vaccination versus infection across gestation has not been well-characterized. To address this question, we measured the cord to maternal nAb ratio across three trimesters to gain a deeper understanding of the degree of neonatal immunity conferred by maternal SARS-CoV-2 infection or vaccination. In the vaccinated cohort, matched cord NT50 values were 1.6-fold higher (*P* = 0.018) compared to maternal NT50 values in mothers vaccinated during the first trimester, and 1.7-fold higher (*P* < 0.0001) during the second trimester notably, suggesting increased transfer efficiency for mothers vaccinated during this time (**Figure 4A**). In third trimester, no significant difference was found between maternal and cord NT50 values. There was no detectable difference between transfer of neutralizing activity against the WT, Alpha, Delta, and Mu variants individually (**Supplemental Figure 3A**). In the infected cohort, cord NT50 values were not observed to be higher than maternal in any trimester; on the contrary, maternal NT50 values were 1.5-fold higher (*P* = 0.0057) than cord NT50 values with third trimester infections (**Figure 4A**), suggesting less efficient of infection-elicited nAb transfer than that after vaccination. Similar to vaccinated mothers, no difference in neutralizing activity against the different strains was noted (**Supplemental Figure 3B**).

**Figure 4.**
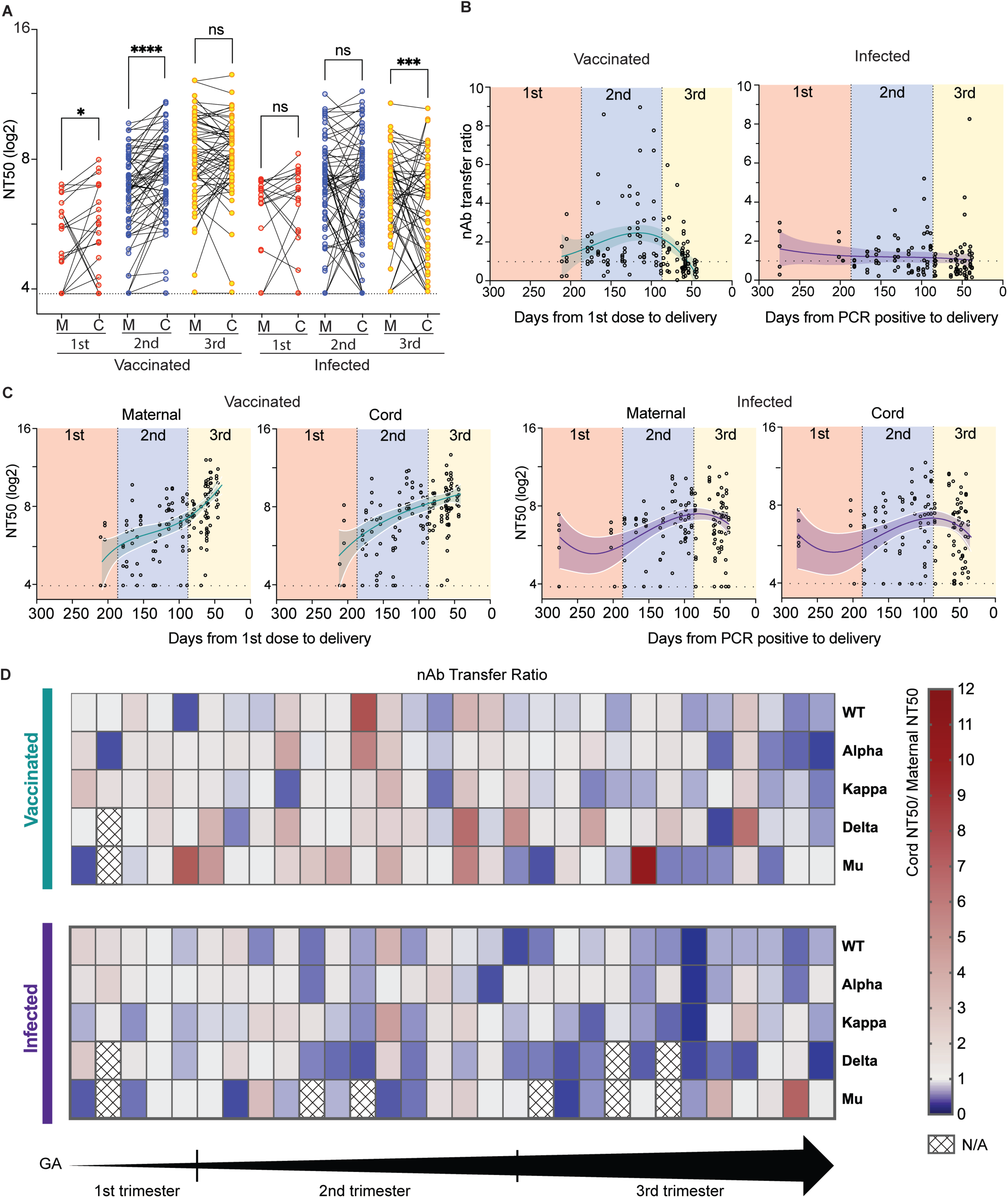
Placental Transfer of SARS-CoV-2 Antibodies. (**A**) The effect of trimester on antibody transfer. The dot plots show NT50 titers for a cohort of vaccinated maternal-cord pairs. Three separate analyses were performed on the trimester at time of first exposure (Red, first trimester; blue, second trimester; yellow, third trimester). Lines connect mother:cord dyads. Significance was determined by the Wilcoxon signed-rank test. (**B**) Maternal-cord transfer ratios (TR) of nAbs were calculated by cord NT50/maternal NT50 to assess efficiency of nAb transfer. nAb TRs are shown by timing on the first vaccine dose or the first positive PCR result. Correlations between the cord to maternal nAb ratio and days from events were analyzed using non-linear regression analysis. (**C**) Maternal and cord NT50 values were shown by timing on the first vaccine dose or the first positive PCR result. (**D**) Patient based nAb TRs for each SARS-CoV-2 strain in both vaccinated and infected patients. All patients were ordered according to increasing gestational age (GA) within their groups. N/A, samples are not enough. \**P* < 0.05; \*\*\**P* < 0.001; \*\*\*\**P* < 0.0001; ns, not significant.

To further evaluate the correlation between *in utero* transfer efficiency of nAb and the timing of exposure, we analyzed maternal:cord nAb transfer ratios (TR) based on the days from exposure to delivery. Interestingly, the regression lines were distinct between the vaccinated and infected group (**Figure 4B**), demonstrating different transfer kinetics of vaccination- and infection-elicited nAbs. We found that the TR was <1 when mothers were vaccinated in the third trimester and less than 60 days prior to delivery. When > 60 days passed from first dose vaccination to delivery, the 73% of the TRs were > 1. Peak TR in our vaccinated cohort was 114 days prior to delivery (TR =2.5), which corresponded to vaccination at ∼ 24 weeks gestational age. Compared to vaccination, infection-induced antibody transfer did not show significant change over gestation. There was no difference noted in transfer efficiency between the two vaccine manufacturers (not shown). When we analyzed the maternal and cord NT50 values based on the days from exposure to delivery, they were increased with the approach to delivery in vaccinated group (**Figure 4C**).

To determine whether differences in nAb activity was explained by lower total IgG levels in patient samples, we analyzed the nAb titers relative to total anti-N and anti-RBD IgG antibodies. Both maternal and cord NT50 titers were significantly correlated with anti-SARS-CoV-2 IgG titers (**Supplemental Figure 3C and 3D**). However, greater variability was observed in the infected group, in which Pearson’s r was 0.5090 (*P* < 0.0001) for maternal plasma and 0.5500 (*P* = 0.002) for cord plasma. In the vaccinated group, the correlation between NT50 and total IgG titers in maternal plasma was stronger than that in cord, with a Pearson’s r of 0.8821 (P < 0.0001) and 0.6365 (P < 0.0001), respectively.

To obtain a deeper understanding regarding the transplacental transfer of maternal nAb among SARS-CoV-2 strains, we performed clustering analysis of TR by participant (**Figure 4D**). In general, the TR was significantly higher in vaccinated cohort (mean = 1.83) compared to infected cohort (mean = 1.15; *P* = 0.0002) (**Supplemental Figure 3E**). TR was not significantly influenced by SARS-CoV-2 strains on its own, however, when a stratified analysis based on SARS-CoV-2 strains was conducted, TR for Delta variant was significantly higher in vaccinated cohort than in infected cohort (*P* = 0.0022) (**Supplemental Figure 3F**).

### Impact of clinical factors on neutralizing activity

We next explored the impact of clinical factors on maternal and cord nAb activity. First, we analyzed fetal sex on nAbs, which has been shown to impact levels of total anti-SARS-CoV-2 IgG (11). The median NT50 for male fetuses was lower than females in maternal plasma after first trimester vaccination (**Figure 5A**). Of note, all pregnancies in our cohort that were infected in the first trimester had male fetuses. In contrast, nAb was higher in vaccinated maternal, and infected maternal and cord plasma from mothers with male fetuses in the second trimester. No significant difference by fetal sex was observed after exposure in the third trimester in all groups.

**Figure 5.**
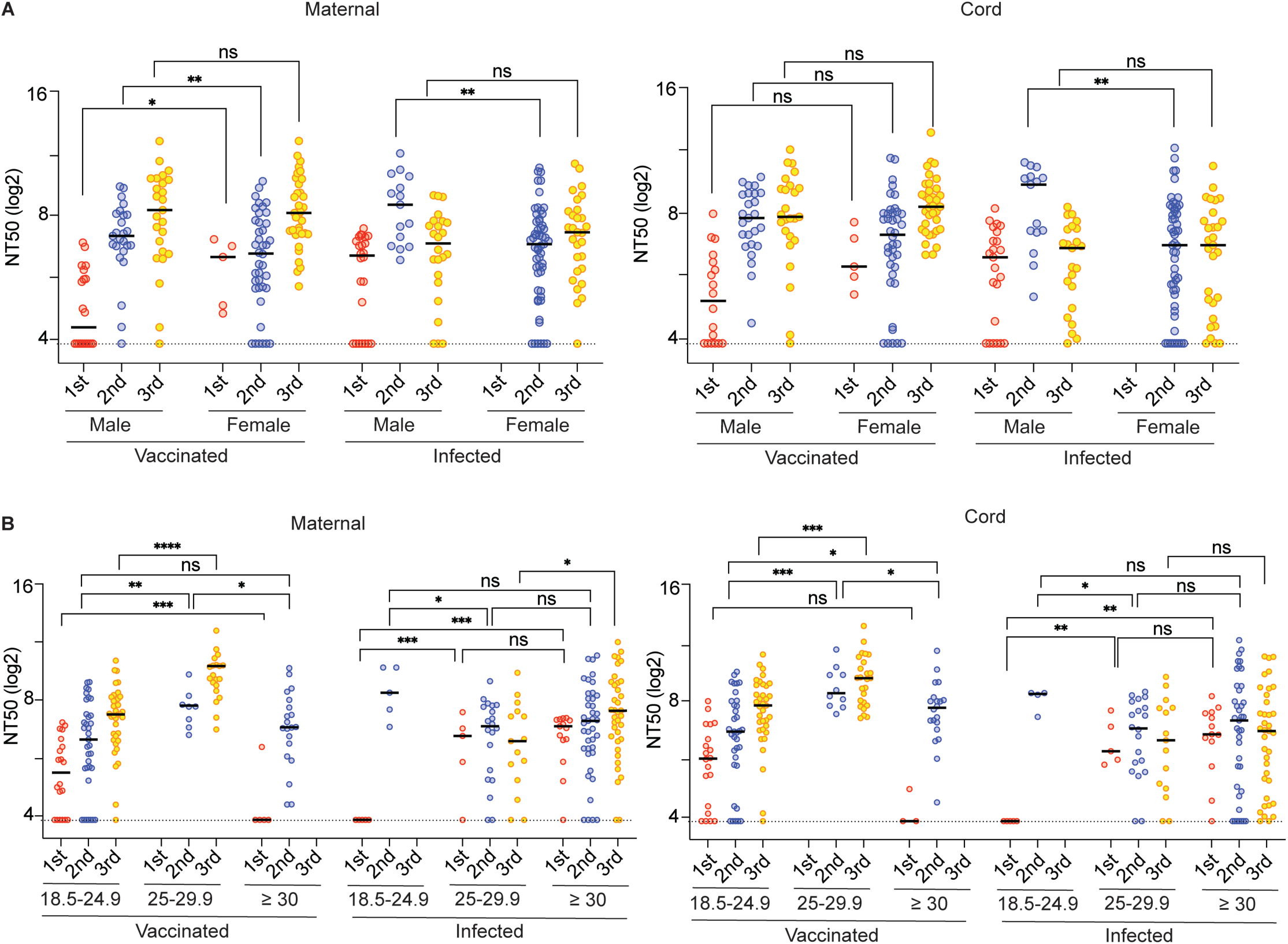
Neutralizing activity and clinical factors. Maternal and cord NT50 values were compared in vaccinated versus infected mothers, stratified by trimester of exposure and by (**A**) Fetal sex and (**B**) Maternal Body Mass Index (BMI). The BMI at the time of delivery was divided into three groups: 18.5 to 24.9, 25 to 29.9, and 30 or more and compared. NT50 values were compared separately for mothers older than 35 years and younger than 35 years. Black lines indicate median. \**P* < 0.05; \*\**P* < 0.01; \*\*\**P* < 0.001 (Mann-Whitney test). Trimester of exposure is indicated by color (Red, first trimester; blue, second trimester; yellow, third trimester).

As obesity is associated with chronic inflammation and dysfunctional immune responses, we next examined the relationship between maternal BMI and neutralizing activity, stratified by trimester. Vaccinated individuals with BMI 25-29.9 at the time of delivery had significantly higher NT50 values in both maternal and cord blood than normal weight (BMI <25) or obese (BMI ≥ 30) individuals (maternal: *P* = 0.001 and *P* = 0.048, cord: *P* < 0.0001 and *P* = 0.47, respectively) (**Figure 5B**). In contrast, infected individuals with BMI 25-29.9 had significantly lower NT50 values in both maternal and cord blood than normal weight individuals, and no statistically significant difference compared to obese individuals in the second trimester. There were not significant differences based on maternal age, with the exception of increased cord NT50s in mothers ≥ 35 years old vaccinated in the second trimester (**Supplemental Figure 4**).

## Discussion

To our knowledge, this is the first study to directly compare nAb activity against five strains of SARS-CoV-2 in gestational age-matched vaccinated and infected maternal-fetal dyads across the entire course of gestation. Maternal vaccination and infection both produced effective nAbs against SARS-CoV-2, although activity against the Kappa, Delta, and Mu variants were reduced compared to wild-type and Alpha. Overall, maternal nAb titers were comparable between vaccination and infection, while cord nAb titers were higher after vaccination than infection, supporting vaccination during pregnancy to benefit newborns as well as mothers. Mothers vaccinated in the third trimester had the highest level of nAb at delivery in maternal and cord plasma. We found that TR was maximal upon vaccination in the second trimester, while the TR of infection-induced nAbs was stable across gestation. However, vaccine-elicited nAb waned gradually throughout pregnancy, as both the titer and transfer ratio are lowest in mothers vaccinated in the first trimester. Interestingly, we show significant variation in nAb response and nAb transfer between patients. Maternal-fetal transfer efficiency was higher in vaccinated mothers versus infected mothers. Finally, our results demonstrate selective transfer of nAbs of differing specificity from mother to baby. These findings are summarized in **Table 2**.

**Table 2.**
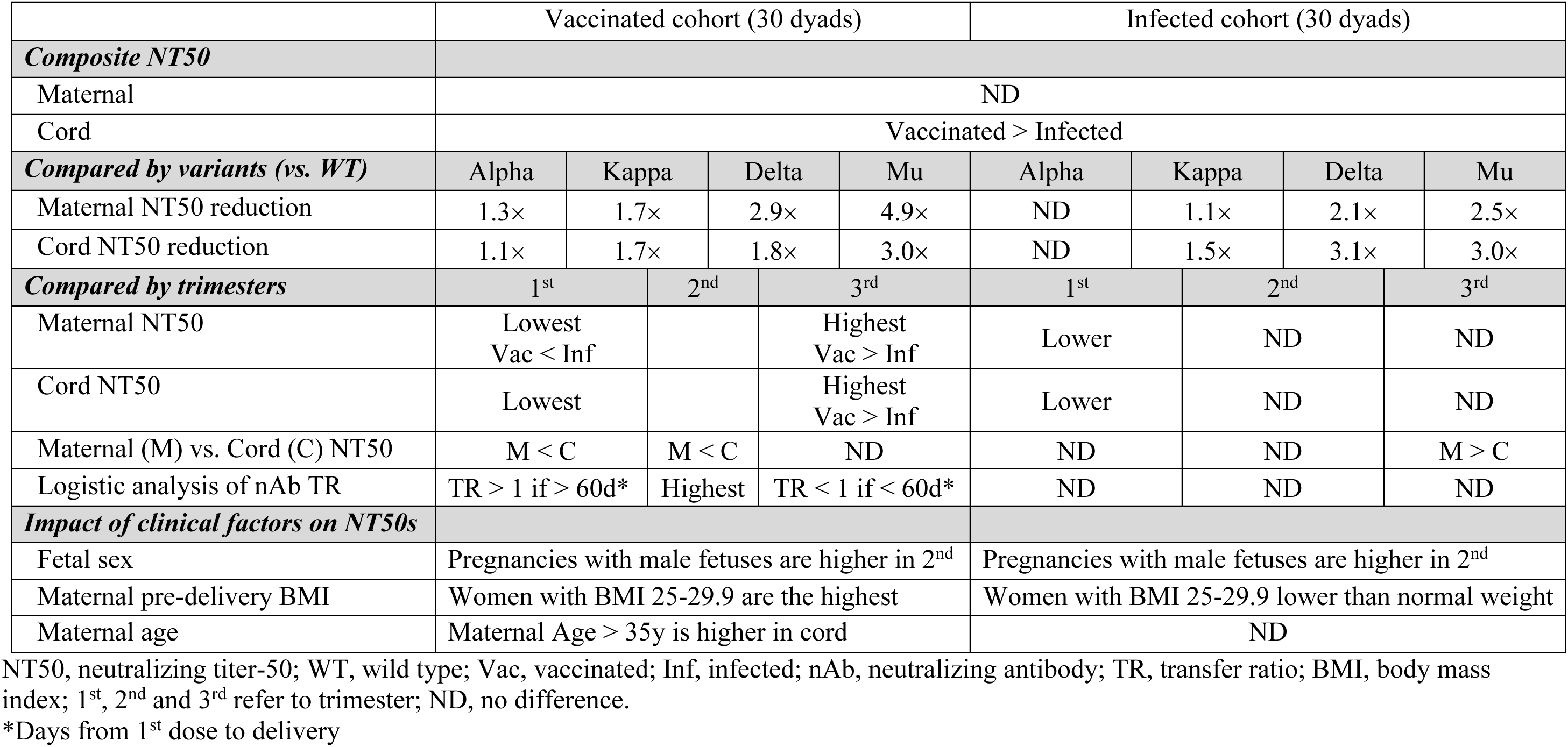
Summary of key findings in this study

Our findings build upon a previous study with a cohort of 30 pregnant women vaccinated in the third trimester showing that mRNA vaccines were immunogenic in pregnancy, but that neutralizing activity against the Alpha and Beta (B.1.351) variants were reduced (34). Here, we provide data for all trimesters of pregnancy and report on nAb activity against the Delta variant, which remains predominant worldwide and accounts for over 99.5% of COVID cases during the 4^th^ wave of the pandemic (35). One of the strengths of this study is the comparison vaccine-induced versus natural immune responses to the Delta variant across pregnancy. We found reductions in nAb titers against the Delta variant in both vaccinated maternal blood and paired infant cord blood, which is consistent with recent reports that the Delta variant is resistant to vaccines in non-pregnant population as compared with the Alpha variant (14, 36). SARS-CoV-2 spike protein RBD is the major target of nAbs, and mutations in the RBD of the Delta variant reduce the neutralization sensitivity of plasma from vaccinated or infected individuals (14).

Furthermore, we have shown that the activity of vaccinated and infection-induced antibody responses against the Mu variant in pregnancy. Our results confirm that in pregnancy, Mu proves to be more resistant to neutralization than all other currently recognized variants (16, 37, 38). The Mu variant can cause cell-to-cell fusion, like the Delta variant, that facilitates escape from the humoral immunity more efficiently than the wildtype virus (37). The Mu also harbors three mutations in the RBD (R346K, E484K, and N501Y) which are not in the Delta variant (27). Neutralizing antibodies targeting the RBD have been divided into four classed based on structural analyses of their epitopes: (class 1) neutralizing antibodies that bind only to “up” RBDs; (class 2) ACE2-blocking neutralizing antibodies that bind both “up” and “down” RBDs; (class 3) neutralizing antibodies that bind outside the ACE2 site and recognize both “up” and “down” RBDs; and (class 4) previously described antibodies that do not block ACE2 and bind only to “up” RBDs (39). The E484K mutation can most strongly disrupt the binding of the class 2 antibodies while L452R is located in the class 3 site (40). In both vaccine and convalescent populations, escape mutants targeting the class 2 antibodies have the most critical impact on impaired neutralization (41). Developing vaccines that elicit polyclonal antibodies with broader epitope specificities may improve the efficacy and longevity of current vaccines. Although the significant and sustained reduction in prevalence of the Mu variant indicates it is unlikely to be predominated over the Delta (42), there is a growing concern that current vaccines may need to be continually modified and updated to against emerging variants with greater properties of immune escape and neutralizing resistance, especially for the vulnerable populations, such as in pregnancy.

Interestingly, nAbs of different variant specificity have differing overall titers and maternal-fetal TRs, which also differs by type of exposure. In our pregnant cohorts, vaccination in the third trimester induced higher nAb levels at delivery, while NT50s were lower with first trimester vaccination compared to infection. The variation of nAb activity between trimesters is more significant in the vaccinated mothers. As reported in the non-pregnant individuals, the range of nAb activity elicited by vaccination is variable compared to that elicited by infection depending on the time after vaccination or infection (43). Consistent with prior studies in non-pregnant individuals (43), we found no differences in nAb specificity across variants between the two mRNA vaccine manufacturers. More studies are needed to understand host factors that determine these variable maternal immune responses and transfer dynamics.

Neonates depend largely on the passive immunity from the maternal antibodies due to functional immaturity of their immune system. Maternal-fetal transfer of IgG against SARS-CoV-2 after infection has been previously reported with TR between 0.72-0.90 (20, 44, 45). However, greater than 85% of infected cases in these studies were diagnosed positive in the third trimester and close to delivery, which may not allow sufficient time for induction of maternal Ab responses and subsequent transfer. Our prior work on a cohort of pregnancies with natural infection, TRs of total IgG was significantly higher when the first maternal positive PCR was 60-180 days before delivery compared with < 60 days (1.2 vs. 0.6, *P* < 0.0001) (33). However, in this study, our infected mothers had stable TRs of nAb across pregnancy. The differences between our findings and that of Song *et al.* (33) may be due to measurement of anti-SARS-CoV-2 total IgG against nucleocapsid protein and RBD in their study versus only anti-RBD antibodies with neutralizing activity here. Our vaccinated mothers showed greatest nAb TRs in the second and early third trimester. We found that nAb TR passes 1.0 if vaccinated >60 days prior to delivery, ranging from 0.04 at 68 days to 9.0 at 114 days (average 2.0). This is similar to the IgG TR in infected mothers previously shown by Song *et al* (33).

Our results also demonstrate selective transfer of antibodies across the placenta. The transport of IgG is thought to be carried out primarily by neonatal Fc receptor (FcRn) in the placental syncytiotrophoblast. FcRn is an atypical Fc gamma (γ) receptor (FcγR) that can bind the IgG Fc region, and has variable affinity for different IgG subclasses (IgG1, IgG2, IgG3 and IgG4) regulated by many factors such as maternal IgG concentrations, disease states, infections and timing of gestation (46–48). Also, Fc glycosylation states influence selective transfer, and reduced fucosylation was found on the Fc domain repertoire of IgG antibodies produced by COVID-19 patients, resulting in enhanced interactions with the activating FcγR (49, 50). It remains to be determined which subclass of IgG is the most effective in neutralizing the SARS-CoV-2, and neonates might not be fully protected from SARS-CoV-2 infection even though amounts of antibodies are detected in cord blood.

Antibody transfer from mother to fetus depends on gestational age and maternal IgG levels. The transport happens in a linear fashion as the pregnancy progresses, with the largest amount transferred in the third trimester (46, 51, 52). Our results are consistent with these findings. Both maternal and cord blood have the highest amount of neutralizing activity in the third trimester. Interestingly, transfer efficiency appears to be greatest in the second trimester vaccination. This may be related to maximizing overall time available for maternal antibody production, as well as time for antibody to transfer and accumulate in the fetal circulation. As expected, the lowest amounts of neutralizing activity were noted at the time of delivery in maternal and cord blood after first trimester vaccination and infection, consistent with the natural decline of overall antibody levels after initial exposure. In whole, our results suggest that pregnant individuals receive their first dose at least 60 days prior to their anticipated delivery date.

As previously reported, effectiveness after only one dose of vaccine is much lower (30.7-48.7%) among persons with the Delta and Alpha variant infection (53). To gain the greatest protection for both mothers and neonates, pregnant women should also receive two doses before delivery. As significantly decreased nAb was observed in the mothers vaccinated in the first trimester, a later booster dose would likely increase the levels of antibodies transferred to the fetus for individuals who vaccinated in early pregnancy. However, further studies are needed to evaluate the dynamics of antibody stimulation from boosters in pregnancy.

Of note, the patient demographics of the two cohorts were different in race and age. This reflects the population-based epidemiology of people that have highest rates of vaccine uptake and COVID-19 infection in the United States (54). This difference also speaks to the fact that there are structural factors (e.g. racism, inequitable access to COVID-19 PPE, testing, and vaccines and quality healthcare) that place pregnant and postpartum women of color at greater risk for being exposed to COVID-19 (5, 6). As vaccine efficacy was generally consistent across subgroups stratified by age, race and ethnicity in general population (55, 56), it is unlikely that these factors would impact the biologic immune response to mRNA vaccine and SARS-CoV-2 in pregnant women. Maternal BMI at delivery was also significantly different between vaccinated and infected cohorts. The majority (27/29, 93%) of our mothers in the infected cohort were overweight, reflecting that obesity is a risk factor for disease severity in individuals with SARS-CoV-2 infection due to reduced respiratory system compliance and impaired innate and adaptive immune responses (57–59). Also, communities of color are at higher risk of obesity due to food apartheid, limited access to healthy foods, food insecurity and communities that do not have high walkability scores or green space. We found that the vaccinated individuals with BMI 25 to 29.9 at delivery can produce the most amount of nAb. Assuming that these patients were of normal BMI (18.5-24.9) prior to pregnancy, the result suggested that the vaccine response is most effective in this group compared with overweight or obese individuals.

A prior study showed that maternal SARS-CoV-2-specific IgG antibody production was reduced and placental antibody transfer was impaired significantly in pregnancies with a male fetus, in which all patients were tested SARS-CoV-2 positivity in the third trimester during admission for delivery (11). Inconsistent with their result, our study showed no significant difference in nAbs by fetal sex after exposure in the third trimester in both groups. Lower nAb activities for male fetuses was only observed in first trimester vaccination, and all pregnancies infected in the first trimester were male fetuses, which suggests pregnancies with male fetuses might be more vulnerable in the first trimester infection due to low nAbs.

There are several limitations and strength of our study. First, we have few patients that were exposed in the first trimester. However, this is balanced against a significant strength of our study in that the participants were matched by timing of vaccine or live viral exposure during pregnancy, allowing direct comparison of vaccine-induced and natural immunity in pregnancy and different timepoints in pregnancy. Second, we do not have sequencing data for the natural infection cohort, and thus do not know which strains caused the infection. However, all infected patients tested positive before January 2021, prior to broad circulation of the Alpha and Delta variants in the United States (60, 61). Presumably, individuals infected with the Delta variant would have higher specific nAb activity against this strain and may also have differing responses to the other variants. Future studies would be helpful to understand the immunogenicity evoked by the Delta, Mu, and other variants in pregnancy. Third, detailed antibody characterization, such as IgG subtyping or fucosylation state, is needed to understand the role of these factors in the dynamics of maternal-fetal antibody transfer. This should be addressed in future studies.

In conclusion, vaccination in pregnancy is highly effective in generating nAbs against all strains of SARS-CoV-2 tested, although activities against the Kappa, Delta, and Mu variants are reduced. Vaccine-induced neutralizing activity is comparable to natural immunity; however, cord levels of nAb are higher after vaccination. There is significant individual variation in antibody responses and antibody transfer from mother to fetus. Vaccination prior to 60 days before delivery is important for transfer of these neutralizing antibodies to the neonate. These results strengthen current recommendations to vaccinate all pregnant people against COVID-19.

## Methods

### Study design and samples collection

Pregnant individuals who had a positive SARS-CoV-2 test by quantitative PCR were enrolled in our study from March 2020 through January 2021 from participating study sites. Pregnant individuals who received an mRNA-based COVID-19 vaccine were enrolled from December 2020 through August 2021 at the University of California San Francisco (UCSF). Thirty vaccinated and 30 infected pregnant women were chosen from these two cohorts to be matched on the approximate gestational age of first vaccine dose and the earliest confirmed SARS-CoV-2 test. Vaccinated participants were fully vaccinated (at least 14 days after the second dose) with the BNT162b2 (17/30) or mRNA-1273 (13/30) vaccine at the time of sample collection. Maternal blood and infant cord blood used were collected at delivery in an EDTA collection tube and were processed within 24 hours. Plasma was isolated from whole blood by centrifugation at 1500rpm for 10min, then aliquoted in the cryovial tubes and stored at -80°C until analysis. Clinical data was abstracted from the medical record.

### Preparation of Pseudotyped Virions

For the preparation of virions, 293T cells are transfected with the spike plasmid followed inoculation with a previously generated working stock of rVSVΔG-rLuc*G (G protein-deficient vesicular stomatitis virus (VSV-G) containing an integrated Renilla Luciferase reporter gene) to generate the pseudotyped rVSVΔG-rLuc*SARS-CoV-2 (62). Pseudotyped virions were generated using spike plasmids harboring mutations found in the WT SARS-CoV-2 spike (Wuhan-Hu-1, GenBank accession number: MN908947.3), the Alpha variant (H69 deletion, V70 deletion, Y144 deletion, N501Y, A570D, D614G, P681H, T716I, S982A, D1118H), the Kappa variant (L452R, E484Q, D614G, P681R, and Q1071H), the Delta variant (T19R, G142D, E156 deletion, F157 deletion, R158G, L452R, T478K, D614G, P681R, and D950N), and the Mu variant (T95I, Y144T, Y145S, 146N insertion, R346K, E484K, N501Y, D614G, P681H, and D950N) (**Figure 1**). The three types of virions were titrated based on the TCID50 method and equalized the infectivity titers.

### Pseudotyped Virion Neutralizing Antibody Assay

To determine the neutralization activity of plasma, pseudotyped virion nAb experiments were performed with Calu-6 epithelial cells (ATCC HTB56) stably expressing human Angiotensin Converting Enzyme 2 (hACE2) (OriGene, RC08442). Twenty-four hours before administration of virion, 2.5×10^4^ Calu-6 hACE2 cells were plated per well of a 96-well plate in 200 μL of complete DMEM. The SARS-CoV-2 spike pseudotyped virions harvested from the supernatant of the 293T cells were assayed for titration and then aliquots mixed for 30 minutes with heat-inactivated plasma samples. Plasma samples were diluted in calcium-free DMEM starting at 1:15 dilution, and then three-fold serial dilutions for 6 final concentrations, in triplicate. The mixtures were then used to infect Calu-6 hACE2 cells and incubated at 37 °C and 5% CO_2_ for 24 hours. At 24 hours post infection the cells were washed once with 1x PBS then 20 μL of lysis buffer was added per well, followed by 100 μL of Renilla Luciferase substrate/buffer (Promega, E2810) according to the manufacturer’s instructions. The plates were read on a luminometer. Results were analyzed by Prism software version 9 (Graph Pad). SARS-CoV-2 neutralization titers (NT50) of the plasma samples were defined as the sample dilution at which a 50% reduction in relative light units was observed relative to the average of the virus control wells. All NT50 titers were calculated as an average of three independent experiments.

### IgG Antibody Measurement

Antibodies to SARS-CoV-2 IgG was measured using the Pylon 3D automated immunoassay system (ET Healthcare, Palo Alto, California) as described previously (63). The antigens are the spike protein receptor-binding domain (RBD) and the nucleocapsid (N) protein. The assay result is expressed in relative fluorescence units. Maternal blood and infant cord blood at delivery were tested.

### Statistics

Differences between three SARS-CoV-2 strains using the same plasma samples were analyzed using the Wilcoxon signed rank test. Differences between the vaccinated group and the infected group were analyzed using the Mann-Whitney test. Correlations between NT50 titers and IgG antibody titers were analyzed using Pearson’s rank test. The variance of maternal NT50 titers among SARS-CoV-2 strains was calculated by deriving Coefficient of Variation (CV) values per participant and performing multiple linear regression to determine if vaccinated/infected cohort or trimester of exposure were significantly associated with CV values (*P* < 0.05). Maternal-cord transfer ratios (TR) of nAbs were calculated by paired cord NT50/maternal NT50 titers to estimate transplacental transfer of nAB. Multiple linear regression analysis was used to determine associations among TR and vaccination/infection cohorts or SARS-CoV-2 strains. Analyses were performed with GraphPad Prism. Heatmaps were created in R studio (pheatmap) and GraphPad Prism.

### Study approval

This study was approved by the institutional review board of the UCSF (IRB# 20-32077), Santa Clara Valley Medical Center (IRB# 20-021), Oregon Health and Sciences University (IRB# STUDY00021569) and Marshall University (IRB# 1662248-1). Written informed consent was obtained from all participants. Demographic characteristics and clinical data were collected through questionnaires and medical record review.

## Data Availability

All data produced in the present work are contained in the manuscript.

## Funding

These studies were supported by the Bill and Melinda Gates Foundation (INV-017035, SLG and VJF), the Marino Family Foundation (MP), the National Institutes of Health (NIAID K23AI127886 (MP) and NIAID K08AI141728 (SLG)), the Krzyzewski Family (APM and SLG), UCSF National Center of Excellence in Women’s Health (SLG, VJF, YA, and VLJ), the Roddenberry Foundation (YM and WCG), Centers for Disease Control and Prevention Foundation (SLG, VJF, YA, and VLJ), the Valley Medical Center Foundation (DS, PJ, and DSR) and individual donors that provided support through our crowdfunding sites https://givingtogether.ucsf.edu/fundraiser/2718761 and https://spark.ucla.edu/project/20775.

## Author contributions

YM, LL, WCG, and SLG designed the study. LL, MP, AGC, YG, VJG, CYL, UJ, MAC, LW, DS, PJ, DSR, BG, JN, MR, LM, IVA and VJF recruited participants and collected samples. YM, LL, AGC, MM, TYT, MMK, BS, JMH, PC, and GRK conducted experiments and acquired data. YM, LL, MP, NO, JR, WCG and SLG analyzed data. YM, LL, NO, and SLG wrote the manuscript. MP, DS, PJ, DSR, IVA, VJF, YA, VLJ, APM, MO, WCG, and SLG provided funding. All authors assisted with editing the manuscript. WCG and SLG supervised the study. Authorship order for the co-first authors was decided by the senior authors.

## Acknowledgements

We thank the mothers and infants who participated in this study.

## FIGURE LEGENDS

**Supplemental Figure 1.** (A) NT50 values for four dyads who were neither vaccinated nor infected and delivered in the same period were included as negative controls. The dotted line indicates the cut-off threshold of this assay. Comparison of the NT50 values by trimester and by five strains in the maternal blood (B) and cord blood (C), respectively. The dot plots show NT50 values. Black bars represent the median of NT50 values. \**P* < 0.05; \*\**P* < 0.01; \*\*\**P* < 0.001; \*\*\*\**P* < 0.0001; ns, not significant (Mann-Whitney test).

**Supplemental Figure 2.** Maternal blood (A) and cord blood (B) coefficient of variation (CV) values were compared between vaccinated and infected cohorts. Maternal blood (C) and cord blood (D) CV values were compared by trimesters. Dashed black bars represent the mean of CV values. ns, not significant (Multiple Linear Regression).

**Supplemental Figure 3.** (A, B) Comparison of neutralizing antibody transfer by trimester and by five strains. (C, D) Correlation between NT50 values and the IgG values. Cord to maternal IgG antibody transfer ratios were plotted using IgG values tested by the Pylon 3D automated immunoassay system in order of the timing on the first vaccine dose (C) or the first positive PCR result (D). (E) Transfer ratios (TRs) for all five strains were compared among vaccinated and infected cohorts. (F) TRs were stratified by SARS-CoV-2 strain and compared among vaccinated and infected cohorts. Dashed black bars represent the mean of TR values. Vac, vaccinated cohort; Inf, infected cohort. \**P* < 0.05; \*\**P* < 0.01; \*\*\**P* < 0.001; ns, not significant (Wilcoxon signed rank test or multiple linear regression).

**Supplemental Figure 4.** Maternal and cord NT50 values were compared in vaccinated versus infected mothers, stratified by trimester of exposure and by maternal age. NT50 values were compared separately for mothers older than 35 years and younger than 35 years. Black lines indicate median. \*\*\**P* < 0.001; ns, not significant (Mann-Whitney test).

## Notes

**Conflict of interest:** The authors have no conflict of interest exists.

### Competing Interest Statement

The authors have declared no competing interest.

### Funding Statement

These studies were supported by the Bill and Melinda Gates Foundation, the Marino Family Foundation, the National Institutes of Health (NIAID K23AI127886 and NIAID K08AI141728), the Krzyzewski Family, UCSF National Center of Excellence in Women's Health, the Roddenberry Foundation, Centers for Disease Control and Prevention Foundation, the Valley Medical Center Foundation and individual donors that provided support through our crowdfunding sites https://givingtogether.ucsf.edu/fundraiser/2718761 and https://spark.ucla.edu/project/20775.

### Author Declarations

This study was approved by the institutional review board of the UCSF (IRB# 20-32077), Santa Clara Valley Medical Center (IRB# 20-021), Oregon Health and Sciences University (IRB# STUDY00021569) and Marshall University (IRB# 1662248-1).

