## Supplemental data for "Neutralizing Antibody Activity Against SARS-CoV-2 Variants in Gestational Age-Matched Mother-Infant Dyads"

### Supplemental Figure 1

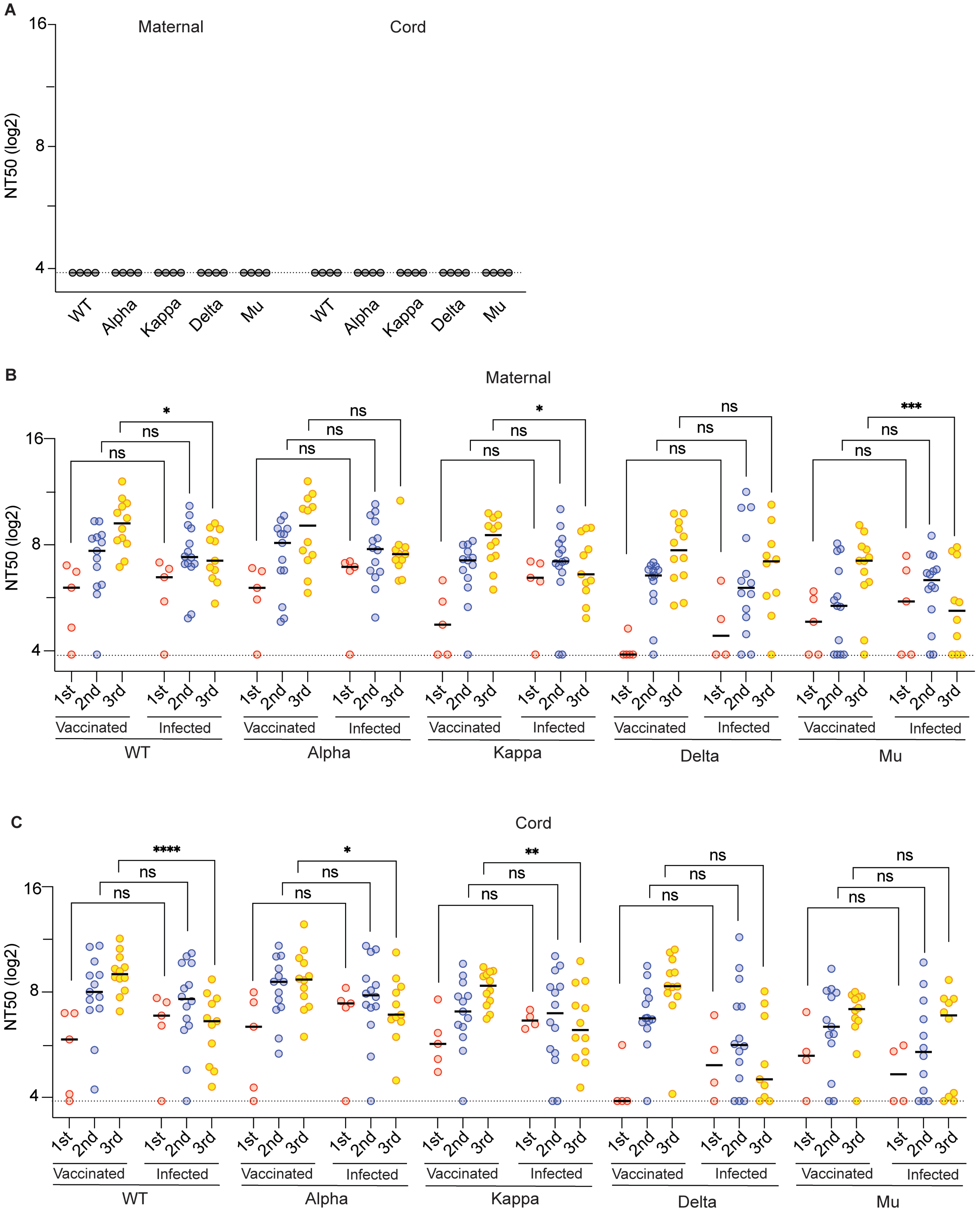


**Supplemental Figure 1.** (**A**) NT50 values for four dyads who were neither vaccinated nor infected and delivered in the same period were included as negative controls. The dotted line indicates the cut-off threshold of this assay. Comparison of the NT50 values by trimester and by five strains in the maternal blood (**B**) and cord blood (**C**), respectively. The dot plots show NT50 values. Black bars represent the median of NT50 values. *P < 0.05; **P < 0.01; ***P < 0.001; ****P < 0.0001; ns, not significant (Mann-Whitney test).

### Supplemental Figure 2

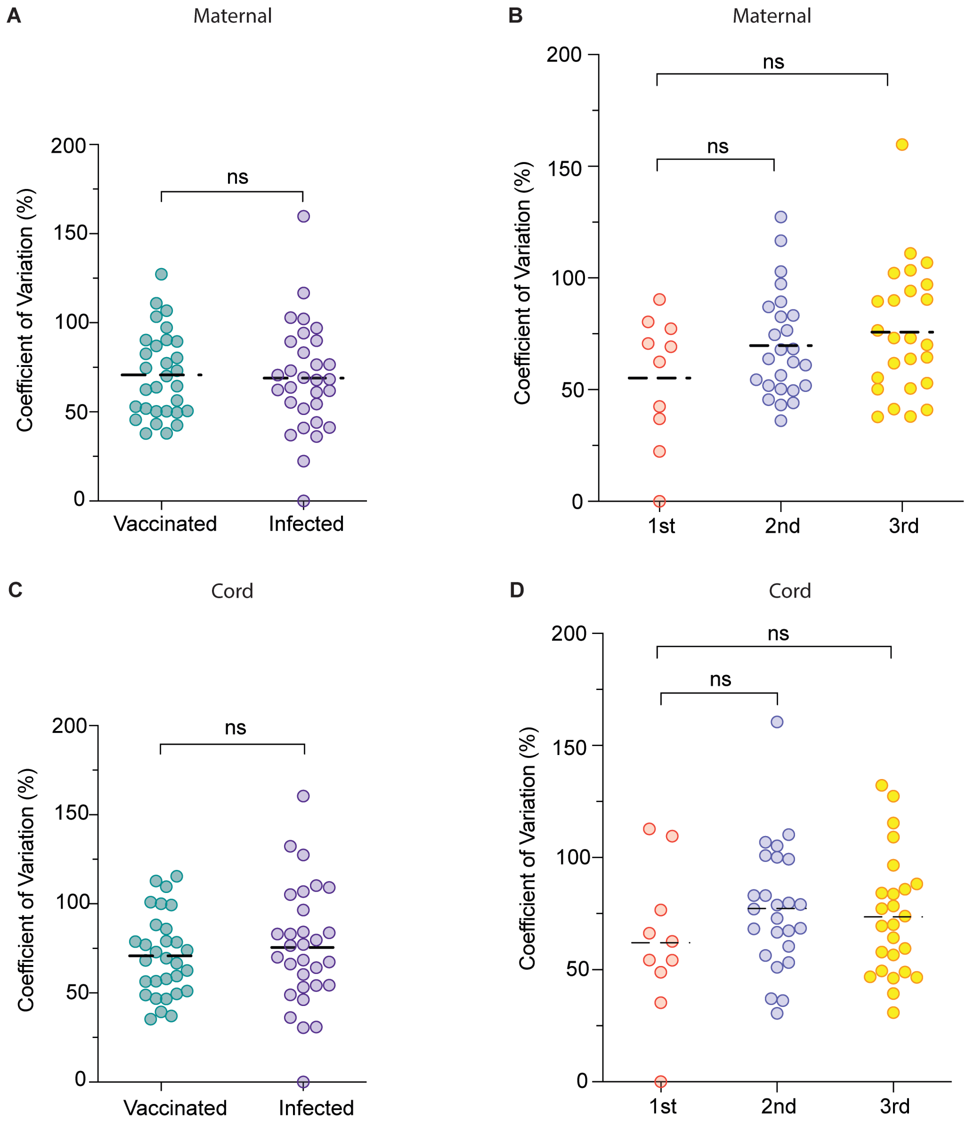


**Supplemental Figure 2.** Maternal blood (**A**) and cord blood (**B**) coefficient of variation (CV) values were compared between vaccinated and infected cohorts. Maternal blood (**C**) and cord blood (**D**) CV values were compared by trimesters. Dashed black bars represent the mean of CV values. ns, not significant (Multiple Linear Regression).

### Supplemental Figure 3

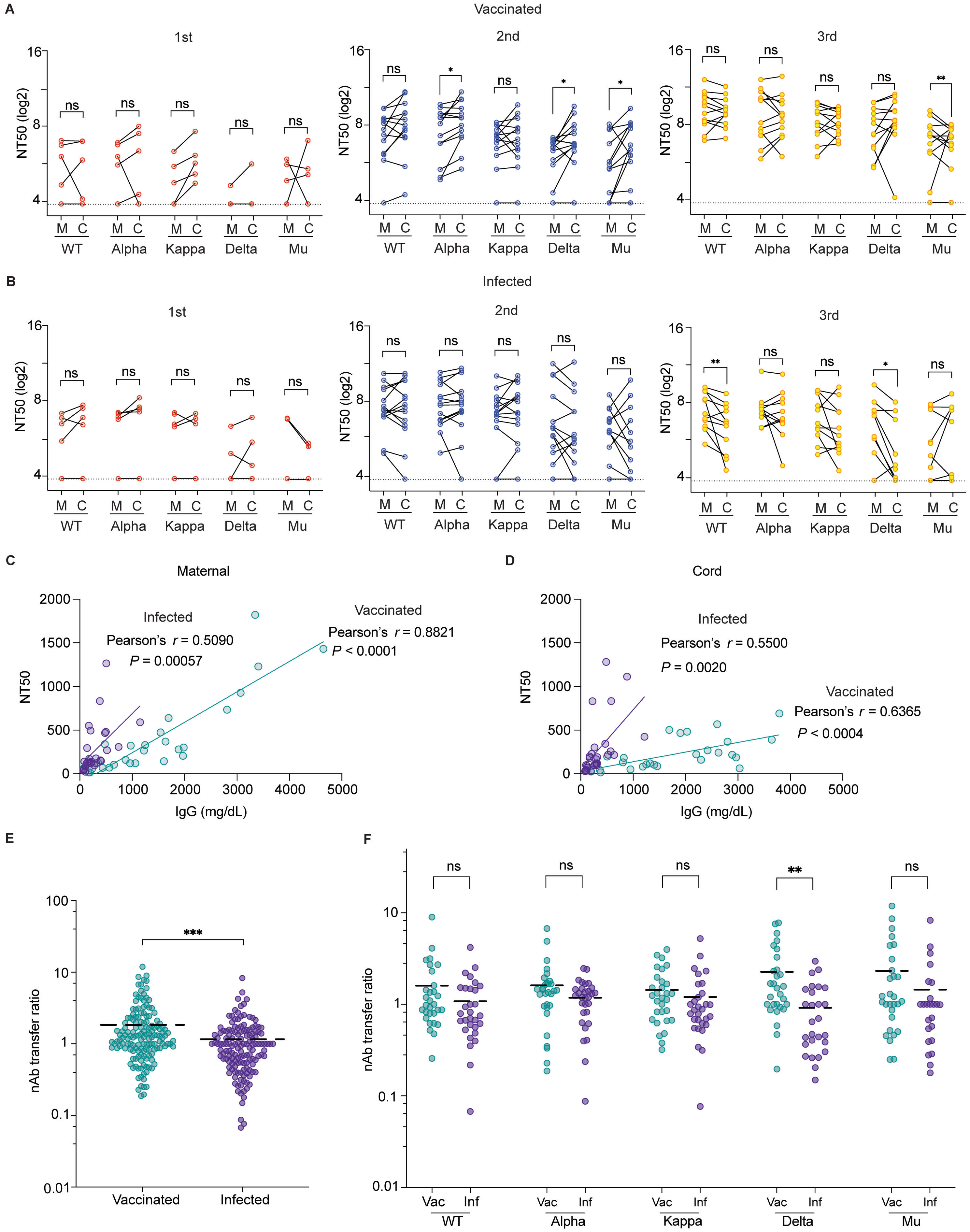


**Supplemental Figure 3.** (**A**, **B**) Comparison of neutralizing antibody transfer by trimester and by five strains. (**C**, **D**) Correlation between NT50 values and the IgG values. Cord to maternal IgG antibody transfer ratios were plotted using IgG values tested by the Pylon 3D automated immunoassay system in order of the timing on the first vaccine dose (C) or the first positive PCR result (D). (**E**) Transfer ratios (TRs) for all five strains were compared among vaccinated and infected cohorts. (**F**) TRs were stratified by SARS-CoV-2 strain and compared among vaccinated and infected cohorts. Dashed black bars represent the mean of TR values. Vac, vaccinated cohort; Inf, infected cohort. *P < 0.05; **P < 0.01; ***P < 0.001; ns, not significant (Wilcoxon signed rank test or multiple linear regression).

### Supplemental Figure 4

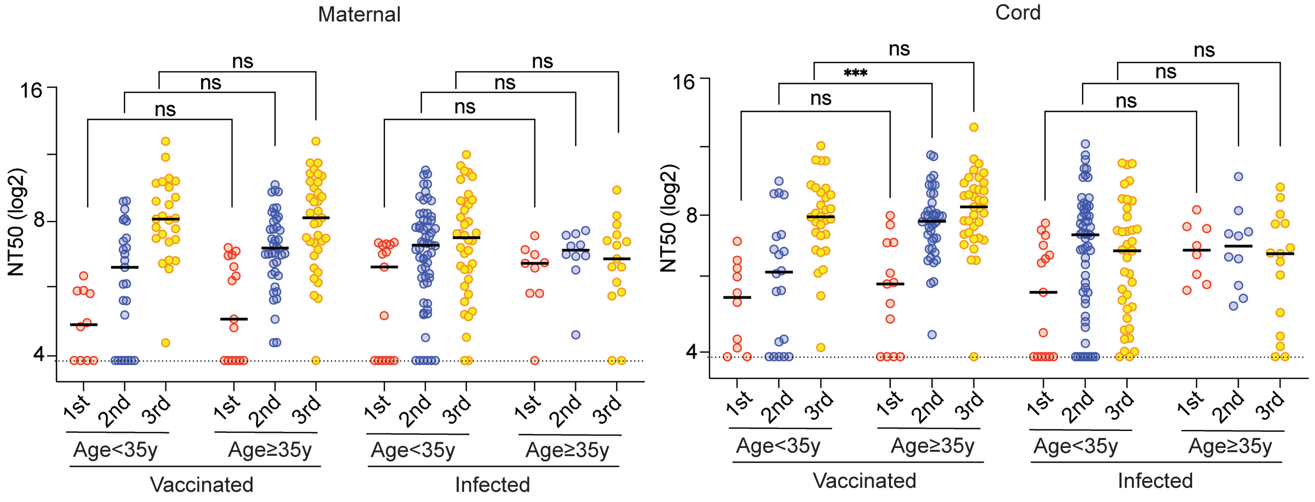


**Supplemental Figure 4.** Maternal and cord NT50 values were compared in vaccinated versus infected mothers, stratified by trimester of exposure and by maternal age. NT50 values were compared separately for mothers older than 35 years and younger than 35 years. Black lines indicate median. ***P < 0.001; ns, not significant (Mann-Whitney test).
